# Heat-based N95 mask decontamination and reuse in a large hospital setting

**DOI:** 10.1101/2020.09.28.20203067

**Authors:** Tushar Vora, Arnab Bhattacharya, Shankar Ghosh, Kiran Gowda, Nandita Dhanaki, Rajul Gala, Nitin Dubey, Vandana Raut, Vikas Kumar Singh, Gagan Prakash, Sarbani Ghosh Laskar, Manju Sengar, Girish Chinnaswamy, C S Pramesh

**Affiliations:** Tata Memorial Hospital/Homi Bhabha National Institute, Tata Memorial Centre, Ernest Borges Road, Mumbai 400012, India; Tata Institute of Fundamental Research/Homi Bhabha National Institute, Homi Bhabha Road, Mumbai 400005, India

**Keywords:** COVID-19, decontamination, N95 masks

## Abstract

The shortage of N95 masks have spurred efforts on developing safe and scientifically-validated decontamination and reuse protocols that are easily scalable and universally applicable even in low-resource settings. We report on the development and implementation of a heat-based N95 mask decontamination system in a large hospital setting (Tata Memorial Hospital, Mumbai, India) with over 8000 N95 masks from about 1400 individual users decontaminated and in reuse till date. We describe the challenges and constraints in choosing a proven, scalable, and easy-to-implement decontamination solution. We discuss the heat treatment and particle filtration efficiency measurement experiments done to validate a decontamination treatment protocol at a target temperature of 70^o^C for a duration of 60 minutes, and the scaling up of this method using a standard hot drying cabinet at the hospital. The logistics of ensuring optimal utilization of the decontamination facility without compromising on basic safety principles are detailed. Our method relies on equipment available in standard hospitals, is simple to set-up, scalable, and can be easily replicated in low-resource settings. We further believe such limited reuse strategies, even in times of abundant N95 mask availability, would not only be cost-saving but also be environmentally responsible in reducing the amount of medical waste.

## Introduction

The use of face masks by health care workers and the public has been universally recognized as a key non-pharmaceutical Intervention in controlling the current COVID-19 pandemic^1^. Filtering facepiece respirator (FFR) masks meeting the N95 (or FFP2) specification are a vital part of personal protective equipment for health care workers. N95 FFRs have traditionally been single-use items typically disposed after short term usage to avoid risk of infection and cross contamination; however, the unprecedented shortages of these masks spurred global efforts at developing scientifically-validated and reproducible methods for N95 decontamination and re-use.^2-5^ Several decontamination methods have been proposed including ultraviolet UV-C irradiation^4,6-8^, microwave oven use^6^, autoclaving and moist/dry heat treatment^4,8-14,15^, and chemical usage (e.g. ethylene oxide^6,16^, hydrogen peroxide vapour^6,7^, chloride dioxide^17^, bleach^6,16^, ethanol^7,15^). However, these need to meet not only stringent criteria for viricidal efficacy,^18,19^ but should also not affect the particle filtration efficiency and fit of the mask,^20^ and not leave chemical residues. Most importantly, the method should be easily implementable on a hospital scale, and ideally be usable even in remote and/or low-resource settings. While several studies have evaluated such decontamination methods, they are primarily laboratory demonstrations of possible protocols, with very few published reports on large-scale hospital implementations. Czubryt et al. recently reported^21^ a heat-based decontamination study with a cohort of only 14 N95 respirator samples. In this report, we document the development and implementation of a heat-based N95 decontamination system at a large hospital (Tata Memorial Hospital (TMH), Mumbai) with over 8000 N95 masks decontaminated and in reuse till date.

### Tata Memorial Centre / Hospital, COVID-19 and N95 masks

Tata Memorial Centre / Hospital (TMC/TMH) is the oldest and largest comprehensive cancer care centre in India. Located in Mumbai, TMH registers more than 75,000 new patients with cancer each year, with the majority coming from distant places. Early on, in March 2020, when the implications of COVID-19 pandemic were beginning to be felt worldwide, a conscious decision was taken by TMC to continue and not compromise cancer care (albeit with extensive adaptive modifications), lest the long-term consequences be much more devastating, and also take care of patients and staff who would invariably, at some point, be exposed to and acquire COVID-19 illness. The details of various strategies employed to fulfil our goal to continue care of cancer with COVID-19 have been reported earlier.^22^ To cater to the need of ∼1500 staff requiring N95 mask usage we anticipated a demand/supply mismatch and started collaborative efforts with Tata Institute of Fundamental Research (TIFR) for optimal selection of N95 masks and developing methods for decontamination and reuse in March 2020 itself.

### Selection of Method for N95 Mask Decontamination and Reuse

We selected the most appropriate, effective and pragmatic method for decontamination of N95 masks on a large scale based on five criteria: 1. Proven viricidal efficacy, 2. No degradation of mask filtration efficacy, 3. Preservation of mask structural integrity and face fit, 4. Lack of chemical residues, and 5. Ease of logistical implementation in a large hospital setting. For the last point, our challenges were not only the large numbers of masks but also the safety of the personnel who carried out the decontamination, and simplicity of returning the decontaminated mask to the same user, in a timely manner. The practical issues in setting up a decontamination system became even more important as India, and Mumbai in particular, went under a very strict lockdown from the end of March, severely constraining the availability of equipment and manpower.

Of the reported decontamination techniques, a review of prior literature and technical summaries published by N95decon consortium (www.n95decon.org), suggested that the suitable methods for our purpose were UV-C irradiation, hydrogen peroxide vapour treatment, and heat-based techniques. Chemical disinfection methods were deemed risky due to residues or uncertain impact on filtration efficiency. Simple procedures like treatment with dilute bleach or alcohol-based solutions would not be acceptable since they remove the electrostatic charge on N95 mask filter media and degrade filtration efficiency.^23^ Implementation of UV-C methods was fraught with difficulties like elimination of shadows, providing uniform illumination of sufficient intensity and most importantly, availability of UV-C germicidal lamps during the lockdown. Large hydrogen peroxide vapour based commercial systems (e.g. the Battelle^24^ or Steris^25^ decontamination systems) were not available in India. Heat-based decontamination methods were hence felt to be most suitable due to easy applicability, availability and accessibility at a fraction of the cost as compared to other methods. With availability of autoclaves and hot drying cabinets in almost all hospital settings, we felt that successful demonstration of this method would make it easier to replicate in similar scenarios.

Heat-based inactivation of pathogens via autoclaving is a standard sterilization method used in hospitals world over. However, some N95 masks have failed fit tests after even one standard autoclave cycle.^1313^ A careful control of temperature, humidity, and decontamination time is required to achieve viral inactivation while maintaining N95 filter performance. Further, the decontamination conditions effective for mask filtration media surfaces may vary from those used in studies on standard media used in experimental conditions. Recent studies have shown that the SARS-COV-2 virus can be inactivated by exposure to temperatures of 70°C for 30-60 minutes depending on the ambient humidity. A heat-treatment time of 60 minutes at 70°C with no added humidity was found to cause >3 log reduction in the SARS-COV-2 virus^7^ though the study was not on N95 mask material. Other reports have shown that compared to dry heat, higher humidity (>50%) required shorter treatment times of ∼30 minutes at 70°C,^9,11^ similar to previous studies on the influenza virus.^26^ Most of these studies have also shown that respirator performance was not degraded after 5 treatment cycles at 75–85°C at 60–90% relative humidity for 30 minutes. Since longer heating times would be more effective in ensuring adequate decontamination (provided they did not degrade mask fit and filtration performance) we explored a treatment regime of 70°C for 60 minutes, the necessary conditions for “dry heat” decontamination, while also providing additional humidity in the oven environment.

### Heat based protocol tested on single masks at TIFR

Experiments were carried out in a laboratory oven with a 45 cm x 45 cm x 40 cm chamber (Trishul Instruments, Mumbai). Apart from the oven temperature control display, temperature and humidity sensors were taped on the test N95 masks to ensure that the actual mask surface conditions could be monitored (See Fig. 1). For this a temperature and humidity meter (based on a Sensirion SHT-31 T/RH sensor, or EL-USB-2 temperature/humidity logger) were used. For the first trial, a 3M 8822 N95 mask (a common mask being used at TMH) was subjected to 5 heating cycles at a target temperature of 70°C for a duration of 60 minutes at the target temperature. The set point was adjusted to achieve a mask surface temperature just above 70°C. For masks kept directly on the oven rack the surface temperature reached the target temperature within 7-10 minutes. For masks kept inside (an open) paper bag, the mask temperature reached the target temperature in about 10-12 minutes time. The temperatures recorded in the 5 cycles was between 70-75°C. While the temperature of the oven could be easily controlled via a PID controller, no in-built humidity control was available. Given the local laboratory conditions (ambient humidity ∼75-80% in Mumbai) the minimum humidity at 70°C was ∼12-15%. Sheets of tissue paper dosed with tap water were kept on the base of the oven and saturated before use. This increased the humidity of the oven to 25-40% during the 60 minute period.

**Fig. 1:**
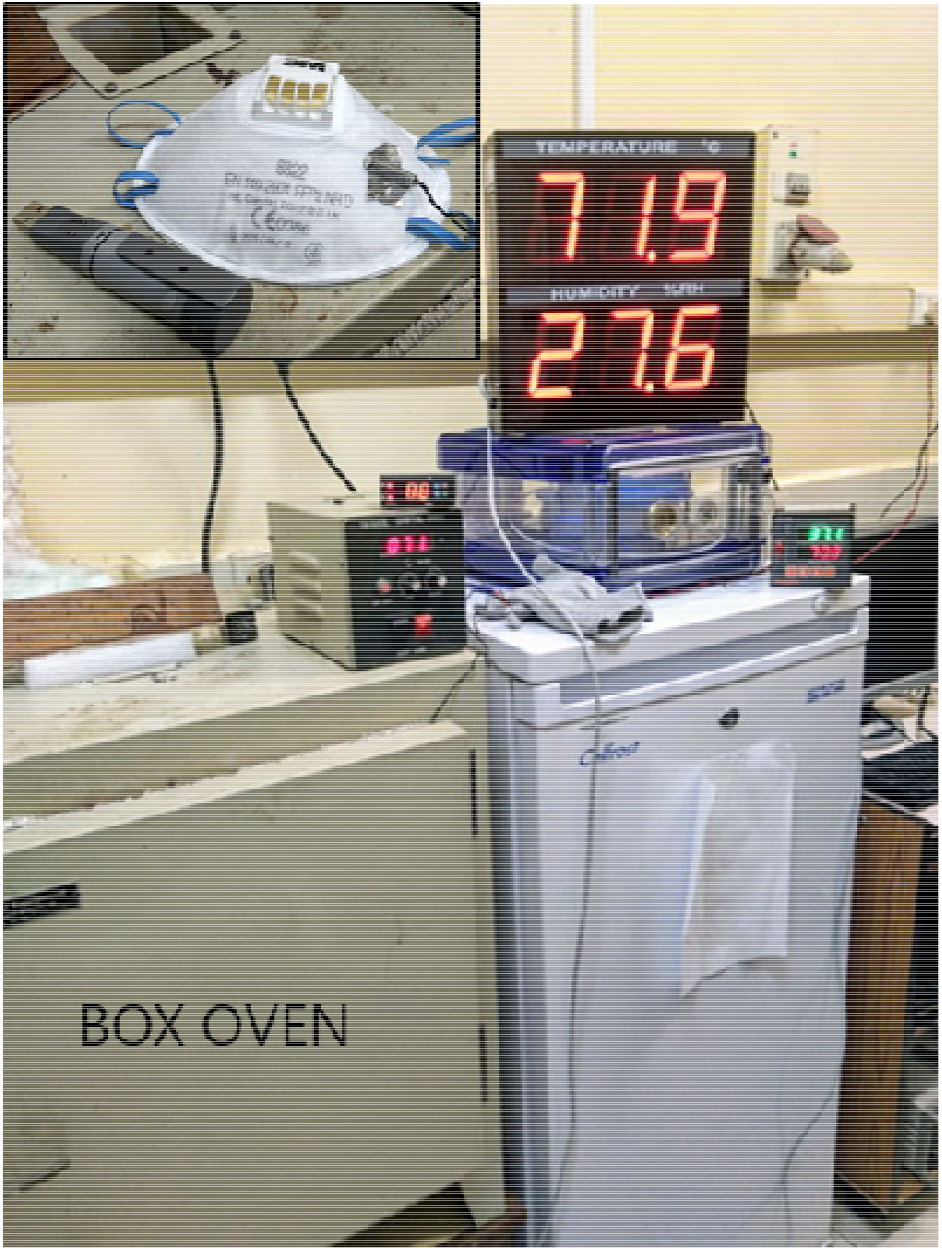
Box oven used for heating experiments at TIFR. Inset: 3M 8822 mask with the temperature/humidity probe stuck on it with conductive metal tape.

Following each heat treatment cycle the particle filtration efficiency (PFE) for 0.3μm sized particles was measured. Because of the COVID-19 pandemic and consequent lockdown, we did not have access to commercial mask filtration test equipment, and the measurements were performed on a home-built setup to measure the PFE using an air-quality monitor as a particle counter. In our test setup air is sucked through the particle counter using a vacuum pump (at ∼10 lpm, similar to human breathing rates) and the throughput of 0.3 μm-sized particles (for N95) measured with and without a mask, and the PFE determined from the ratio of these counts. A stable background of 0.3 μm particles was created using normal saline (0.9% NaCl solution) in a nebulizer to make a fine aerosol. Details of the mask filtration test system are available on a GitHub repository.^27^ Since the entire mask was tested, the sides of the mask were taped to the holder to ensure that there was no leakage of particles from the sides.

PFE data for the pristine mask and after the 5 heat treatment cycles are shown in Fig. 2(a). We also carried out the same heat treatment for 3 cycles on a Venus 4400 N95 mask. Further, a 5 cycle heat treatment study was also performed on two types of surgical masks commonly available in Indian hospital settings, data are shown in Fig. 2(b). In all cases, we did not observe any change in the filtration efficiency compared to the pristine masks. Additionally other masks commonly used in hospitals in India (Venus 4200, Chinese KN95 masks, Magnum 95001) were also subjected to similar heat treatment cycles with no measureable degradation in filtration efficiency.

**Fig. 2:**
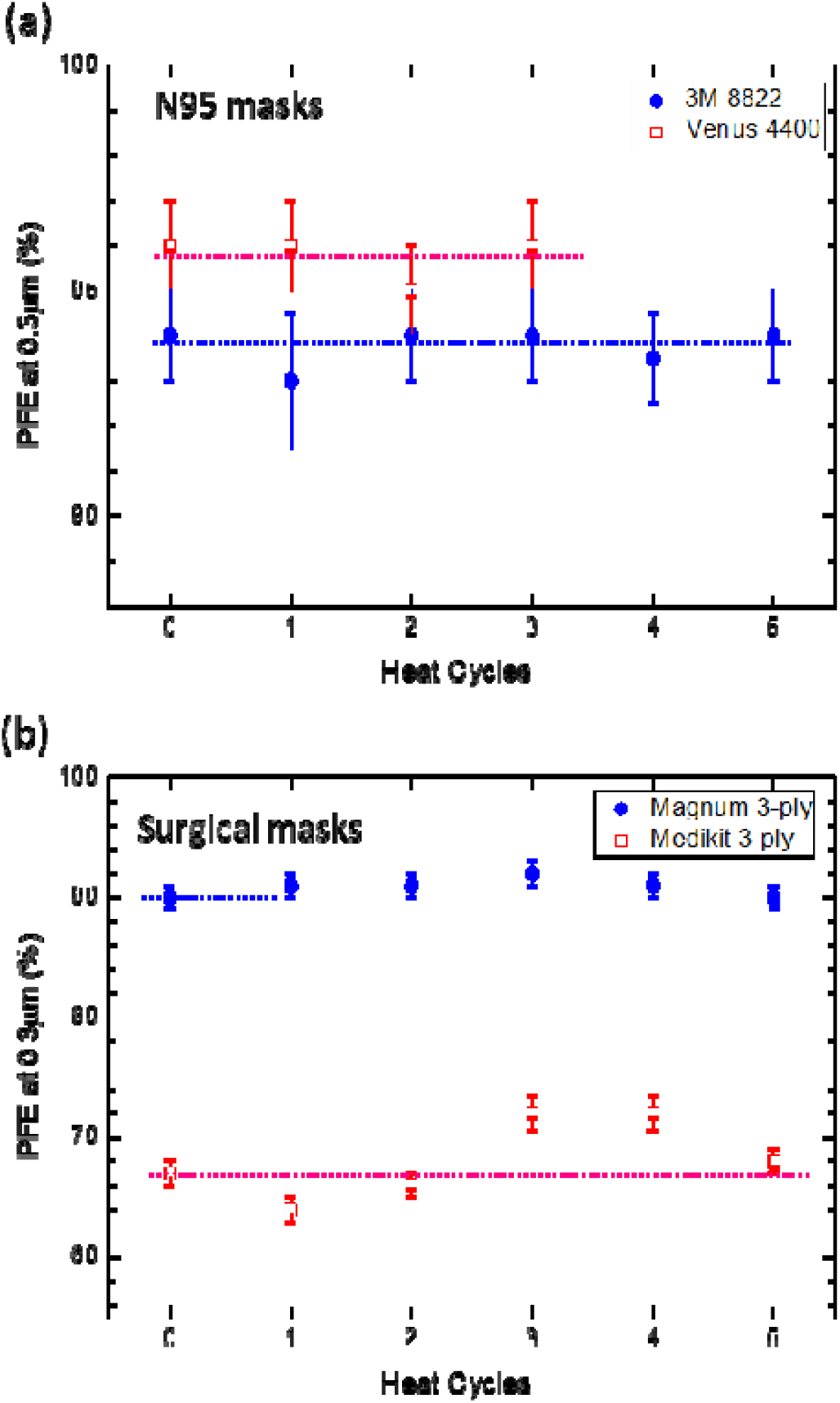
Particle filtration efficiency values (at 0.3μm size) for different heat treatment cycles for (a) N95 masks and (b) surgical masks. Within the limits of our measurement no degradation in filtration efficiency was seen.

To check if the heating cycles had any effect on the microstructure of the fibers, electron microscopy studies were carried out on the filter layers of the N95 mask. Small samples were punched out of both a pristine and the heat-treated mask, the various layers separated out using a tweezer, and examined in a Zeiss EVO scanning electron microscope. The comparative images of the layers shown in Fig. 3 suggest that there is no change in the microstructure before and after the 5 heat treatment cycles, corroborating the filtration efficiency measurements.

**Fig. 3:**
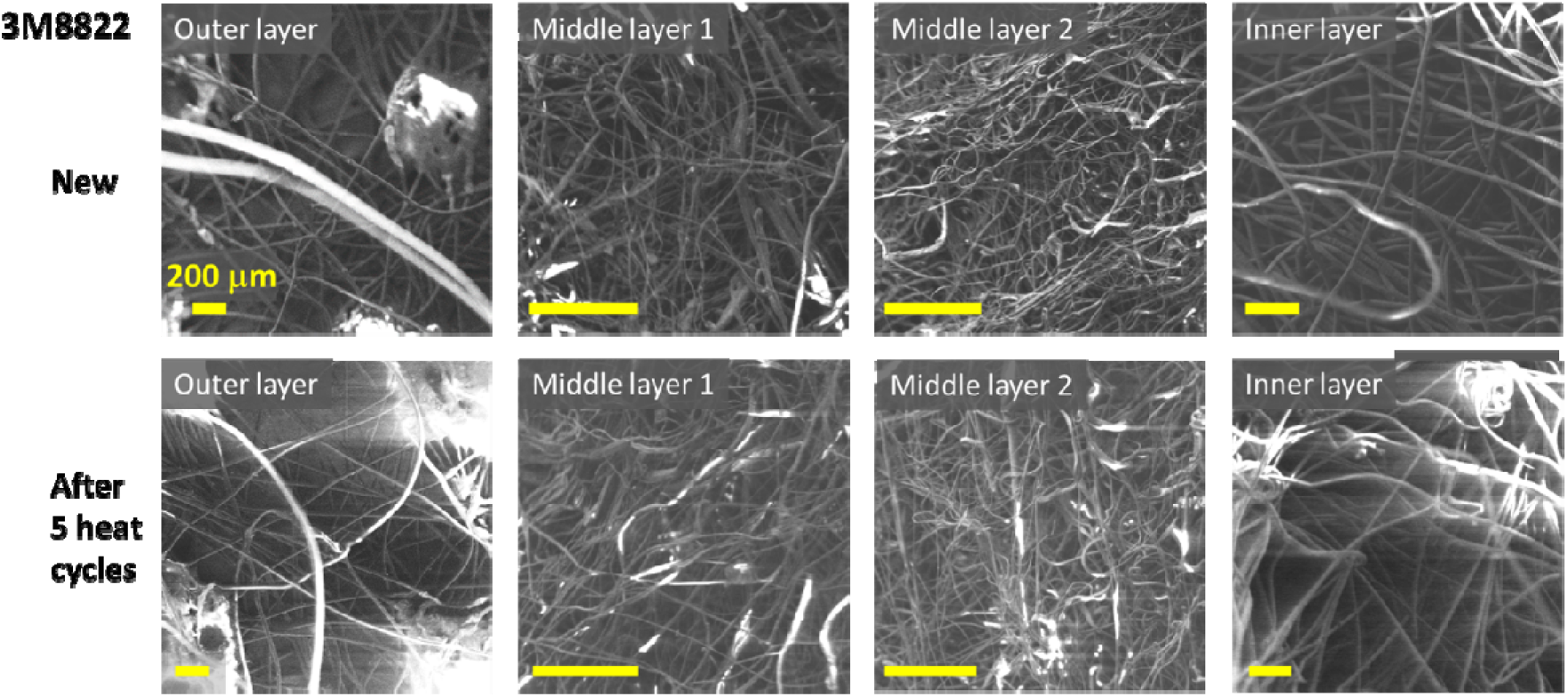
Scanning electron microscopy images of the different layers of a 3M8822 mask. The top row shows images of a pristine mask, the bottom row is the mask after 5 heating cycles (70°C 1 hour). The regular pattern of large white spots in the outer layer are impressions from the thermal calendering process used to make the spun-bond non-woven fabrics. All scale bars are 200μm.

## Scale up and Implementation at a Pan-Hospital level

After selection of dry heat method for decontamination, and having tested the structural and functional integrity post decontamination, we were faced with the bigger challenge of scaling the process to cater to a multi-department, large-hospital setting. With more than 2000 N95 respirator users, requiring decontamination in a periodic manner, we developed several practical but innovative approaches to address this problem of scaling while optimally balancing reuse and safety.

An appropriate heat drying cabinet (Axyos Technologies) was identified in the Central Sterilization Services Department (CSSD) of TMH that could maintain the required ∼70 C temperature uniformly in all parts of the dryer. The dryer has two vertical halves each approximately 180 cm tall, 60 cm wide and 90 cm deep, with grilled racks that can be placed at desired heights. Apart from the in-built temperature displays for the left and right halves of the drying cabinet, two external temperature sensors and temperature/ humidity sensors were used to monitor the temperature at various locations in the cabinet. First, the temperature uniformity of the drying cabinet around a 70°C setpoint was evaluated. While the uniformity across a single rack was within ±1 °C, there was a greater temperature variation from top to bottom of ∼5°C, as expected due to convective heat flow.

Previous implementations of heat-decontamination for N95 masks, e.g. Anderegg et al.^9^, have suggested keeping masks in individual sealed plastic boxes, each with a small tissue paper with a carefully dosed amount of water using a micropipette, to ensure similar humidity levels in each box. An important difference between this and our work is that we use open-top paper bags^*^ to hold the masks for a decontamination, as opposed to sealed boxes. While this necessarily implies that we cannot ensure that each mask is subject to the exact same conditions of temperature and humidity, we believe our method greatly simplifies the logistics of handling a large number of masks. The use of paper bags serves three extremely important functions: 1. Individual bags are labelled by users making it easy to trace and return the masks to the respective users; 2. The CSSD and Decon Team never had to come in direct contact with the N95 masks, minimizing the chance for contamination; and 3. The open top paper bag naturally allowed the mask to “air out” in storage.

To ensure that the N95 masks inside the paper bags reached the target temperature, a set of calibration experiments were carried out. Temperature probes were attached to dummy masks in paper bags, which were kept at various locations in the drying cabinet. Based on these experiments the heating setpoint was kept at 75 °C to ensure that the mask temperature at any location would be ≥ 70 °C. While each rack could hold 16 paper bags, to ensure adequate airflow a maximum of 12 bags were loaded per rack to allow for adequate airflow. In this configuration a maximum of 96 masks could be loaded for decontamination in one heating cycle. A picture of the drying cabinet with a full load of masks is shown in Fig. 4.

**Fig. 4:**
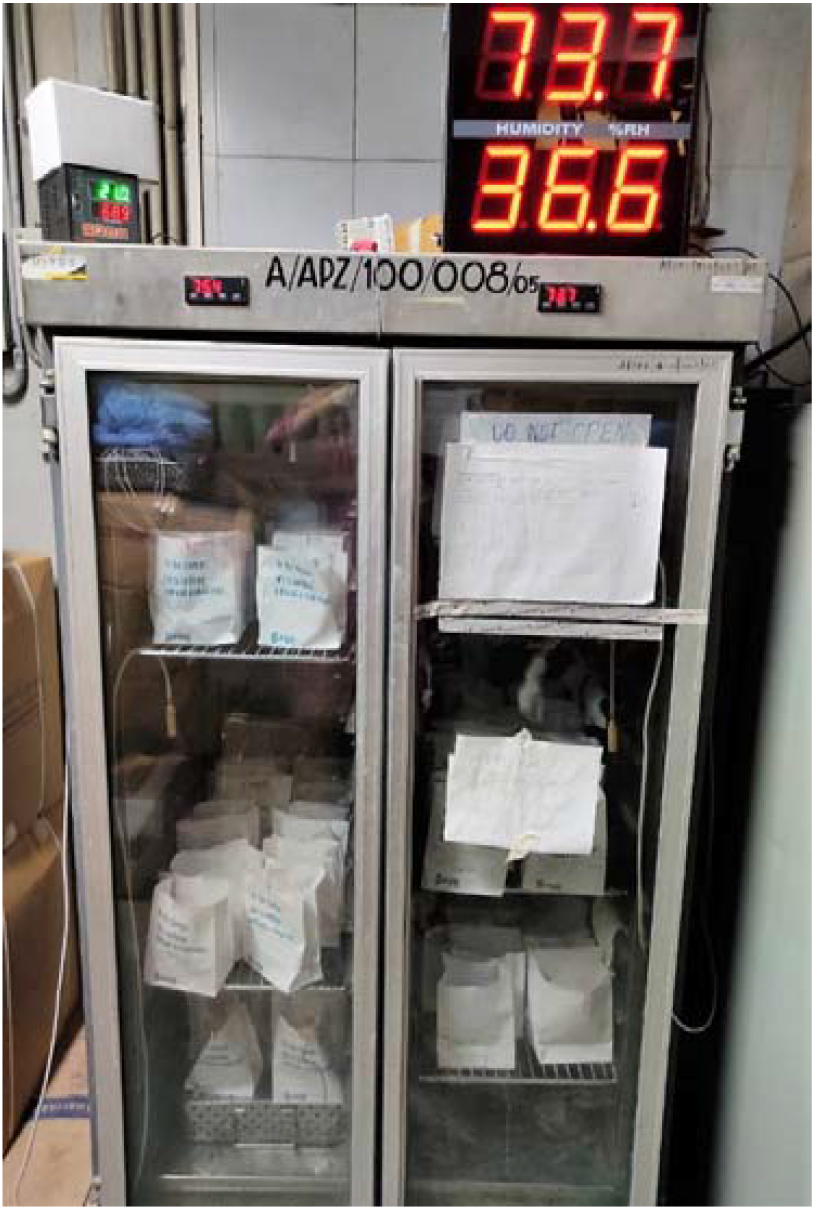
The Axyos drying cabinet used for the heat-based decontamination process at the hospital, with a load of N95 masks kept in open-top paper bags.

Opening the door of the cabinet to load the entire set of paper bags caused a temperature drop of ∼10°C, and it took ∼15 minutes for the temperature to re-stabilize to the setpoint value. The one hour cycle time was calculated after the temperature had reached a stable value. A wet towel was kept at the bottom of the cabinet before the start of each run, resulting in a humidity level of typically 35% at the beginning of each run, decreasing to ∼10-14% at the end of the run. While the 70°C one hour conditions ensured decontamination as per previous dry heat studies, humidity was an additional attempt to increase the efficacy.

## Logistics of decontamination

Hospital-wide staff were divided into 65 subunits (departments, disease management groups, surgical, medical, radiation, radiology, nursing, etc.) depending on location of the facility, number of personnel in each subunit and expected requirements for N95 mask decontamination (Table 1). From each of these 65 units, a unit-coordinator was selected to ensure smooth functioning. To ensure optimal utilization of the decontamination facility without compromising on safety, weekday slots were meticuolously planned among the 65 units to prevent overcrowding, ensure safe distancing and smooth flow of deposit/collection of N95 masks from designated locations. Detailed SOPs were put into place with instructions and expected compliance at user level and unit-coordinator level and the decontamination team (Table 2).

**Table 1.**
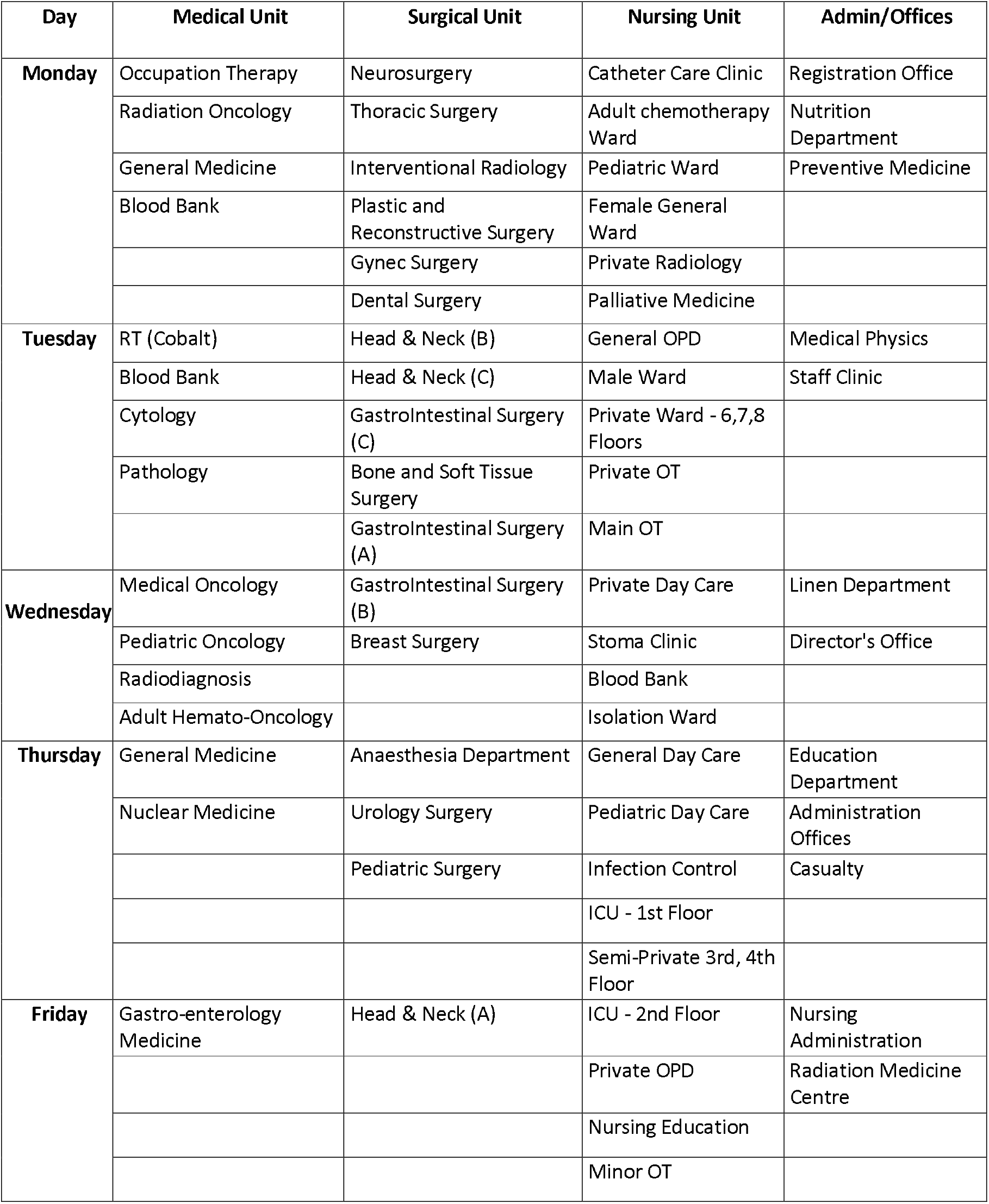
Division of Hospital-wide staff into units and subunits to facilitate even distribution during weekdays and optimal utilization of N95 mask decontamination facility

**Table 2.** SOPs for Individual Mask User, Unit Coordinator and CSSD/Mask Decon Team

|  |
| --- |
| **Individual Mask User SOPs**<br>*Please ensure the following SOPs are followed by each member of the unit.*<br>1. **Identify** the N95 masks allocated to you that needs to be decontaminated (**already used up to 5 times at 96 hours intervals**)<br>2. **Label** the given **paper bag** with your Name, Department and Institute Code Number and put each **individual mask into separate paper bag (ONE MASK PER BAG, Do not put more than one mask in one bag)**<br>3. **Do not close/staple the bag. Keep the bags open.**<br>4. The paper bags (with the masks in them) will undergo the **decontamination procedure** with **strict adherence to technical SOPs** by the Decon Team and the CSSD staff.<br>5. **Collect** the decontaminated masks (in paper bags) from your unit-coordinator.<br>6. After collection of the decontaminated masks - **PLEASE label the mask (not the paper bag)** with permanent marker. (The same mask can be decontaminated up to 3-5 times) |
| **Unit Coordinator SOPs**<br>1. Please Coordinate within your unit for collecting all N95 masks (each in individual labelled paper bags) requiring decontamination according to the protocol mentioned above.<br>2. Please coordinate to send the collected masks, by **single person** (on trolley or in box), to **the designated location (Room No. XXX, near CSSD), between 9 AM and 10 AM** on the designated day – so as to avoid overcrowding in the area.<br>3. Please send a single person for **collection of sterilized masks at 3 PM on the same day** for the entire unit, from the same designated room. He/She can then distribute to individual unit members. |
| **CSSD and Mask Decon Team SOPs (simplified)**<br>1. A day prior, **notify and remind** subunits to deposit N95 masks to be decontaminated between 9 and 10 AM<br>2. **Prime the Heat Dryer Oven** at 8 AM - check temperature, probes, space and electric supply<br>3. At the time of collection, **document** the unit, department, names of mask users, number of masks deposited by each coordinator<br>4. After collection of paper bags (with mask in them), take them to the Heat Dryer Oven and place on shelves with **adequate air flow** guaranteed between individual masks<br>5. Place **temperature probes** on maximum shelves possible to record temperature on each shelf<br>6. **Close and ensure the seal of doors of the oven. Wait till temperature across all probes reaches > 70°C.** Note the time and keep the doors sealed for at least 60 minutes<br>7. After the cycle, note the time and temperature and take out paper bags (with masks in them) and take them to the designated point for recollection by the unit coordinator<br>8. **DO NOT TOUCH the masks directly at any point.** Handle the paper bags NOT the masks<br>9. Handover to respective unit coordinators between 3 and 4 PM |
| **General Instructions**<br>1. The decontamination process is designed to kill viruses and most bacteria NOT fungal spores.<br>2. Filtration and structural integrity of 3M, Venus and Filtra masks have been tested post heat decontamination method in-house. Rest of the makes have not been tested but literature suggests other N95 masks can also be decontaminated using dry heat methods.<br>3. Do not submit structurally damaged masks for decontamination.<br>4. If the masks show signs of fungal patches (stains) do not submit for decontamination. Please discard them and get new masks.<br>5. Do not apply any solutions or reagents to the masks to be decontaminated. It may compromise the decontamination process as well as damage the masks.<br>6. Multiple usage of masks by itself induces a peculiar smell. Decontamination process does not augment or eliminate the smell. |

After setting up the above logistics, the decontamination facility of TMH became functional on May 28, 2020. Till Sept 22, 2020 8193 N95 masks of ∼1400 individual users, have undergone decontamination and are in reuse. Apart from the monetary cost savings, this endeavour has demonstrated the feasibility of such an exercise on a large-scale pan-hospital setting. The most common reason for rejection of user-deposited masks before decontamination was visible stains and fungal mold on the fabric. Physical distortion of structure was seen in 8 N95 masks after decontamination process. The damage appeared due to solvent treatment of mask before heat treatment but could not be proven.

## Conclusions

We have developed an easy and scalable method of heat-based N95 FFR decontamination and demonstrated its deployment in a large hospital setting. Our method of subjecting the masks to target heating conditions of temperature of 70°C for a duration of 1 hour using a hot drying cabinet found in most hospitals can be replicated widely with no additional expenditure. Except for viricidal experiments (for which we have depended on established literature), filtration and structural integrity checks were done in-house. A key to successful utilization of such a decontamination facility is the optimization of the logistics of implementation across the hospital, from developing SOPs for individual users and decontamination system operators to scheduling appropriate time-slots for various sections. While these decontamination procedures were developed out of sheer necessity in crisis periods of N95 mask shortages, we believe that such re-use processes, which significantly reduce medical waste generated, may cause a shift to a more environmentally-responsible usage of N95 masks even under normal circumstances of abundant availability.

## Data Availability

None

## Acknowledgements

We thank Emroj Hossain and Mahesh Gokhale for technical assistance and several useful discussions, and acknowledge the support of the TIFR electron microscopy group for SEM imaging. The work was supported through internal grants from TIFR and TMH.

## Footnotes

* These are standard “sickness bags” – paper bags with an internal waterproof coating. The bags were heated to ∼90°C for 2 hours to ensure that there was no deterioration of the paper bag material.

